# An increase in willingness to vaccinate against COVID-19 in the US between October 2020 and February 2021: longitudinal evidence from the Understanding America Study

**DOI:** 10.1101/2021.03.04.21252918

**Authors:** Michael Daly, Andrew Jones, Eric Robinson

## Abstract

**Background:** Recent evidence suggests that willingness to vaccinate against COVID-19 has been declining throughout the pandemic and is low among ethnic minority groups.

**Methods:** Observational study using a nationally representative longitudinal sample (N =7,840) from the Understanding America Study (UAS). Changes in the percentage of respondents willing to vaccinate, undecided, or intending to refuse a COVID-19 vaccine were examined over 20 survey waves from April 1 2020 to February 15 2021.

**Results:** After a sharp decline in willingness to vaccinate against COVID-19 between April and October 2020 (from 74.0% to 52.7%), willingness to vaccinate increased by 8.1% (*p* <.001) to 60.8% between October 2020 and February 2021. A significant increase in willingness to vaccinate was observed across all demographic groups examined and Black (15.6% increase) and Hispanic participants (12.1% increase) showed particularly large changes.

**Conclusions:** Willingness to vaccinate against COVID-19 increased in the US from October 2020 to February 2021.

**Funding statement:** N/A

## 1. Introduction

As of February 28, 2021, the COVID-19 pandemic had led to over 2.6 million deaths worldwide including over half a million deaths in the United States.^1^ Since the beginning of the pandemic on March 11, 2020, governments across the globe have enacted stringent lockdown and distancing restrictions to curb the spread of the virus. As a result, the pandemic has been linked to adverse societal outcomes including reduced national economic output, declines in population mental health, and disruption to the provision of healthcare.^2,3^ In this context, the rapid development of effective vaccines against COVID-19 offers a path towards population immunity and a return to normal life.

Simulation studies suggest three quarters of the population may need to be vaccinated to achieve herd immunity.^4^ However, recent large-scale reviews indicate that willingness to vaccinate has been declining throughout the pandemic and may be insufficient to ensure widespread vaccination.^5,6^ For instance, in the U.S. approximately half of adults indicated they were willing to be vaccinated against COVID-19 in October 2020, down from over 70% in April, 2020.^7^

By mid-November 2020, results of the Pfizer-Biontech COVID-19 vaccine Phase 3 clinical trial were announced suggesting the vaccine was well tolerated and 95% effective against the virus.^8^ The Pfizer-Biontech vaccine was subsequently approved by the U.S. FDA for emergency use. This was followed by approval of two additional COVID-19 vaccines by companies Moderna and Johnson & Johnson in December 2020 and February 2021 respectively.^9,10^ As of February 28, 2021, nations around the globe were accelerating their vaccination programmes led by Israel where 54.5% of the population have received at least one dose of a COVID-19 vaccine.^11^

The widely publicized emergence of safe and effective vaccines against COVID-19 and the high-profile early successes of mass rollout programmes may have prompted an increase in willingness to vaccinate. To evaluate this possibility, this research examined the percentage of the U.S. population willing to vaccinate against COVID-19 across 20 waves of the Understanding America Study (UAS), a nationally representative longitudinal study of U.S. adults tracked continuously throughout the pandemic. Given concerns about high levels of vaccine hesitancy among ethnic minority groups in the U.S. and elsewhere^5,12,13^ this study also aimed to ascertain whether willingness to vaccinate has increased among Black and Hispanic groups in recent months.

## 2. Methods

### 2.1. Study design and participants

Participants were drawn from the Understanding America Study, a probability-based nationally representative internet-panel of U.S. adults aged 18 and over. The sample was recruited via address-based sampling from the US postal addresses and participants without internet access were provided with internet-connected tablet computers. Details of the UAS survey administration and sampling methodology can found elsewhere .^14,15^

In this study, 8547 eligible UAS participants were invited to take part in biweekly surveys throughout the COVID-19 pandemic. In total, 7,840 participants provided 120,903 survey responses (15.4 surveys on average) across 20 waves of data collection conducted between April 1 2020 and February 15 2021. Participants were assigned a specific day over the course of each two-week survey period to complete the survey providing a continuous account of willingness to vaccinate over this period. The number of participants included in each survey wave and the dates of data collection can be seen in Table S1. The UAS was approved by the University of Southern California human subjects committee internal review board (IRB) and informed consent was obtained from all subjects.

### 2.2. Measures

In each wave of the survey participants were asked how likely they were to get vaccinated against the coronavirus once a vaccine is available. Responses were categorized as either: (1) undecided (responses of ‘unsure’), (2) unwilling to vaccinate (responses of somewhat or very *unlikely* to vaccinate), or (3) willing to vaccinate (responses of somewhat or very *likely* to vaccinate). From December 23 2020 participants were asked if they had received a COVID-19 vaccine. Those who indicated they had received at least one dose of a COVID-19 vaccine were included in the willing to vaccinate group.

Trends in willingness to vaccinate were examined across a set of demographic subgroups based on participant age (18-39, 40-59, ≥60 years), sex (male, female), race/ethnicity (White, Hispanic, Black, Other race/ethnicity), college degree (vs. none), household income (≤$50,00, ≥$50,000 per annum), and chronic health conditions (present vs not present). To assess the presence of chronic health conditions participants indicated whether they had been diagnosed with the following conditions: diabetes, cancer, heart disease, kidney disease, asthma, chronic lung disease, an autoimmune disease.

### 2.3. Statistical analysis

National trends in the public’s willingness to vaccinate against COVID-19 were examined across 20 waves of the UAS conducted between April 1 2020 and February 15 2021. Multinomial logistic regression analysis with cluster robust standard errors were first conducted. The Stata margins postestimation command was then used to estimate changes in the predicted probabilities that participants were willing to vaccinate, undecided, or unwilling to vaccinate at each time-point. Predicted probabilities were multiplied by 100 to represent percentage point changes.

Our initial analyses confirmed that willingness to vaccinate declined from April to October 2020 and increased after this point (see Figure 1). We therefore focused our in-depth analysis of trends in willingness to vaccinate by demographic groups on changes from April 1-14 to the September 30-October 13 wave (hereafter October 2020) and between this point and the January 20-February 15 2021 survey wave (hereafter February 2021). All analyses were adjusted for demographic characteristics and incorporated the UAS sampling weights to adjust for unequal probabilities of selection into the survey and align each survey wave with the distribution of demographic characteristics of the US population.^16^

**Figure 1.**
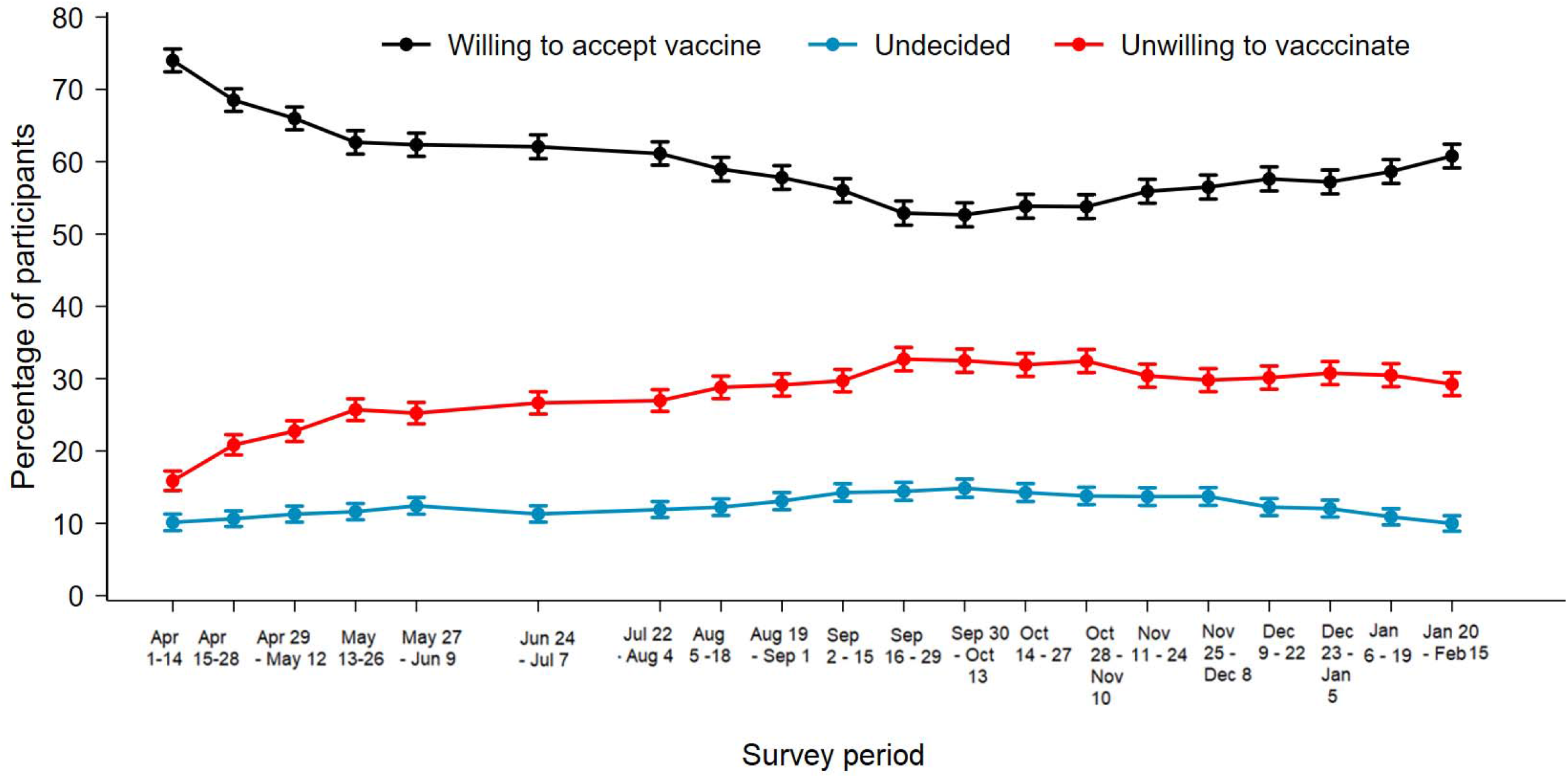
Changes in vaccination intentions across 20 waves of the Understanding America Study conducted between April 1 2020 and February 15 2021. Graph is based on an analysis of 120,903 observations on 7,840 participants and estimates adjust for differences in demographic characteristics between survey waves. 95% confidence intervals are presented.

## 3. Results

### 3.1. Sample characteristics

Sample characteristics are displayed in Table 1. The participants were aged 48.8 (SD =16.6) years on average, 51.5% were female and 66% were White, 16.2% Hispanic, 11.9% Black, and 6% Other race/ethnicity and 33.9% reported being diagnosed with a chronic health condition. On average across all time-points examined 59.5% of the sample indicated they were willing to receive the COVID-19 vaccine when available, 12.4% of participants indicated they were undecided, and 28.1% indicated they were unwilling accept the vaccine.

**Table 1.** Characteristics of participants in the Understanding American Study (UAS; N = 7,840; Observations = 120,903).

| Characteristic | Mean (SD) or % |
| --- | --- |
| Age, years | 48.8 (16.6) |
| Age group (%) |  |
| 18-39 | 37.4 |
| 40-59 | 32.3 |
| 60+ | 30.3 |
| Sex (%) |  |
| Male | 48.5 |
| Female | 51.5 |
| Race/ethnicity (%) |  |
| White | 66.0 |
| Hispanic | 16.2 |
| Black | 11.9 |
| Other race/<br>ethnicity | 6.0 |
| College degree (%) |  |
| No | 64.8 |
| Yes | 35.2 |
| Household income level (%) |  |
| < \$50,000 | 44.7 |
| ≥ \$50,000 | 55.3 |
| Chronic condition<sup>a</sup> (%) |  |
| No | 66.1 |
| Yes | 33.9 |

Weighted estimates are presented.
<sup>a</sup> Diagnosed with any of the following: diabetes, cancer, heart disease, kidney disease, asthma, chronic lung disease, an autoimmune disease.

### 3.2. National trends in willingness to vaccinate

Trends in vaccination intentions between April 2020 and February 2021 are shown in Figure 1. There was a large drop in the percentage of participants willing to accept the vaccine from 73.8% at the beginning of April to 52.8% October 2020 that was observed in the overall sample and across the demographic subgroups examined (see Table 2). At this point willingness to vaccinate was lowest among Black participants (47.1%). From October 2020 willingness to vaccinate increased and 60.9% of adults were willing be vaccinated by February 2021. This group included 10.5% of participants who reported having received at least one dose of a COVID-19 vaccine by the February 2021 survey wave of the UAS.

**Table 2.** Vaccination intentions in the Understanding America Study (UAS) in April and October 2020 and in February 2021 in the overall sample and demographic subgroups.

|  | Willing to accept<br>vaccine |  |  | Unwilling to accept<br>vaccine |  |  | Undecided |  |  |
| --- | --- | --- | --- | --- | --- | --- | --- | --- | --- |
| Survey month | Apr<br>20<sup>a</sup> | Oct<br>20<sup>a</sup> | Feb<br>21<sup>a</sup> | Apr<br>20<sup>a</sup> | Oct<br>20<sup>a</sup> | Feb<br>21<sup>a</sup> | Apr<br>20<sup>a</sup> | Oct<br>20<sup>a</sup> | Feb<br>21<sup>a</sup> |
|  | % | % | % | % | % | % | % | % | % |
| Overall sample | 73.8 | 52.8 | 60.9 | 16.0 | 32.4 | 29.1 | 10.2 | 14.8 | 10.0 |
| Age group |  |  |  |  |  |  |  |  |  |
| 18-39 | 69.6 | 48.2 | 54.0 | 18.9 | 36.0 | 35.3 | 11.5 | 15.8 | 10.7 |
| 40-59 | 72.0 | 48.3 | 58.3 | 17.1 | 35.5 | 30.0 | 10.9 | 16.2 | 11.7 |
| 60+ | 81.0 | 63.2 | 72.0 | 11.1 | 24.8 | 20.8 | 8.0 | 12.1 | 7.2 |
| Sex |  |  |  |  |  |  |  |  |  |
| Male | 79.3 | 59.2 | 65.6 | 13.6 | 28.5 | 26.5 | 7.2 | 12.3 | 7.9 |
| Female | 68.6 | 46.7 | 56.4 | 18.3 | 36.1 | 31.6 | 13.1 | 17.2 | 12.0 |
| Race/ethnicity |  |  |  |  |  |  |  |  |  |
| White | 77.3 | 56.4 | 62.0 | 14.0 | 30.2 | 29.8 | 8.7 | 13.5 | 8.2 |
| Hispanic | 72.8 | 46.9 | 59.9 | 17.2 | 37.7 | 29.5 | 10.1 | 15.3 | 10.6 |
| Black | 48.9 | 31.9 | 47.1 | 29.1 | 44.2 | 32.6 | 22.0 | 24.0 | 20.3 |
| Other race/<br>ethnicity | 87.6 | 69.6 | 78.9 | 8.7 | 20.1 | 14.1 | 3.7 | 10.3 | 7.1 |
| College degree |  |  |  |  |  |  |  |  |  |
| No | 68.1 | 44.4 | 52.9 | 19.1 | 36.9 | 34.1 | 12.8 | 18.7 | 13.0 |
| Yes | 84.8 | 67.9 | 75.6 | 10.0 | 24.3 | 20.1 | 5.3 | 7.8 | 4.4 |
| Household<br>income level |  |  |  |  |  |  |  |  |  |
| < \$50,000 | 66.7 | 43.8 | 51.7 | 18.2 | 36.5 | 32.3 | 15.1 | 19.7 | 16.0 |
| ≥ \$50,000 | 79.5 | 60.0 | 68.3 | 14.2 | 29.2 | 26.6 | 6.3 | 10.8 | 5.1 |
| Chronic<br>condition |  |  |  |  |  |  |  |  |  |
| No | 71.3 | 51.3 | 54.1 | 18.3 | 34.1 | 34.7 | 10.4 | 14.6 | 11.2 |
| Yes | 78.6 | 55.6 | 65.5 | 11.5 | 29.3 | 24.9 | 9.9 | 15.2 | 9.6 |

Note: Weighted willingness to vaccinate estimates are presented.
<sup>a</sup>Survey dates examined were Apr = April 1-14, 2020, Oct = September 30 – October 13 2020,
and Feb = January 20 - February 15, 2021.

In February 2021 29% of participants indicated they were unwilling to accept the COVID-19 vaccine and this figure was particularly high among young adults, Black participants, those without chronic conditions and lower income and education groups (see Table 2).

### 3.3. Regression analyses of changes in willingness to vaccinate between April 2020 and February 2021

Marginal effects computed from a multinomial logistic regression confirmed that the percentage of participants willing to accept the vaccine declined significantly by 21.3% (95% CI[19.4, 23.3], *p*<.001) from 74% to 52.7% between April and October 2020. This significant decline was observed across demographic subgroups (see Table 3) and was explained by an increase in the percentage of participants reporting they are undecided (4.7% increase, 95% CI[3.1, 6.3], *p*<.001) or would refuse the COVID-19 vaccine (16.6% increase, 95% CI[14.8, 18.4], *p*<.001).

**Table 3.**
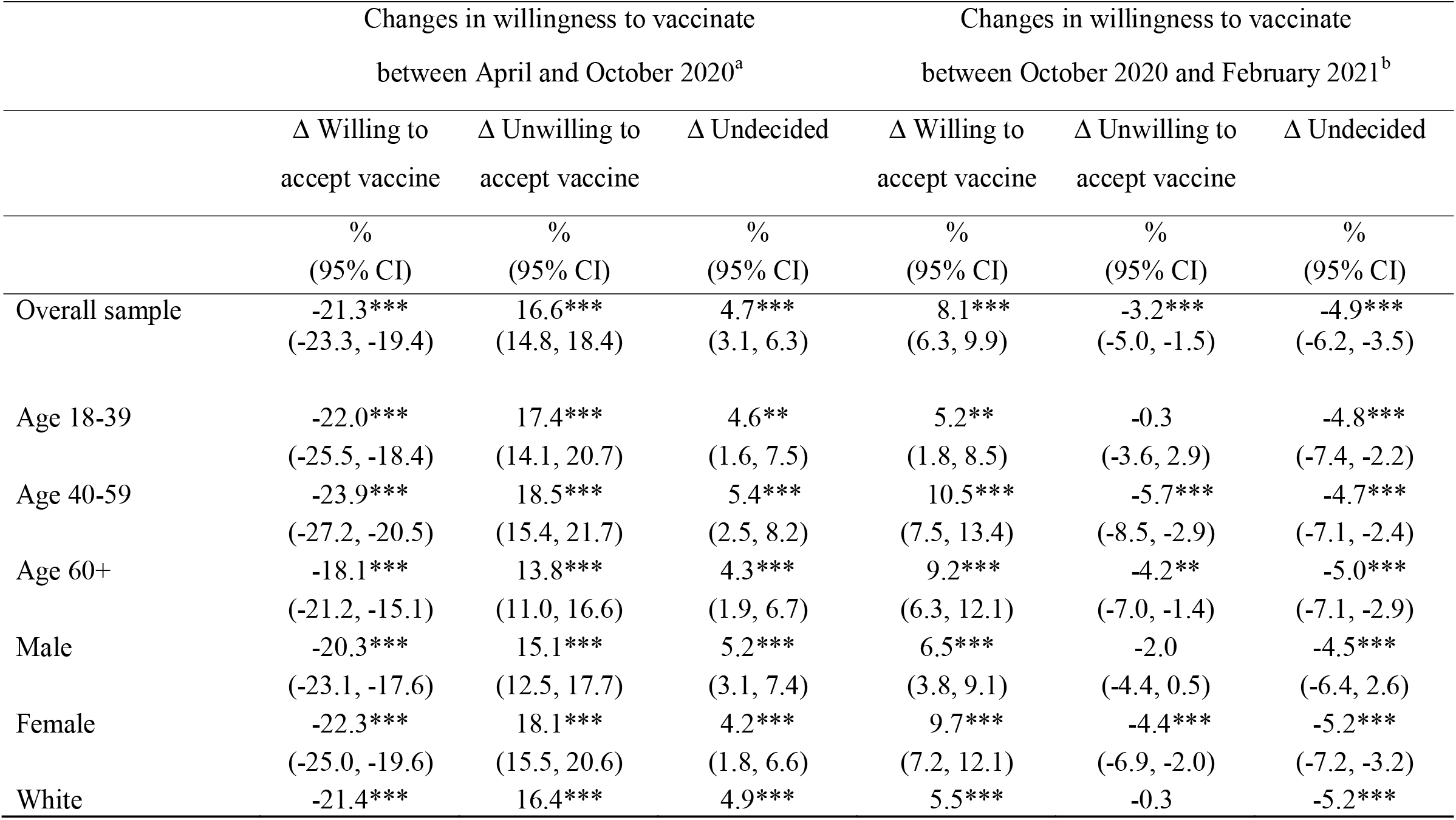
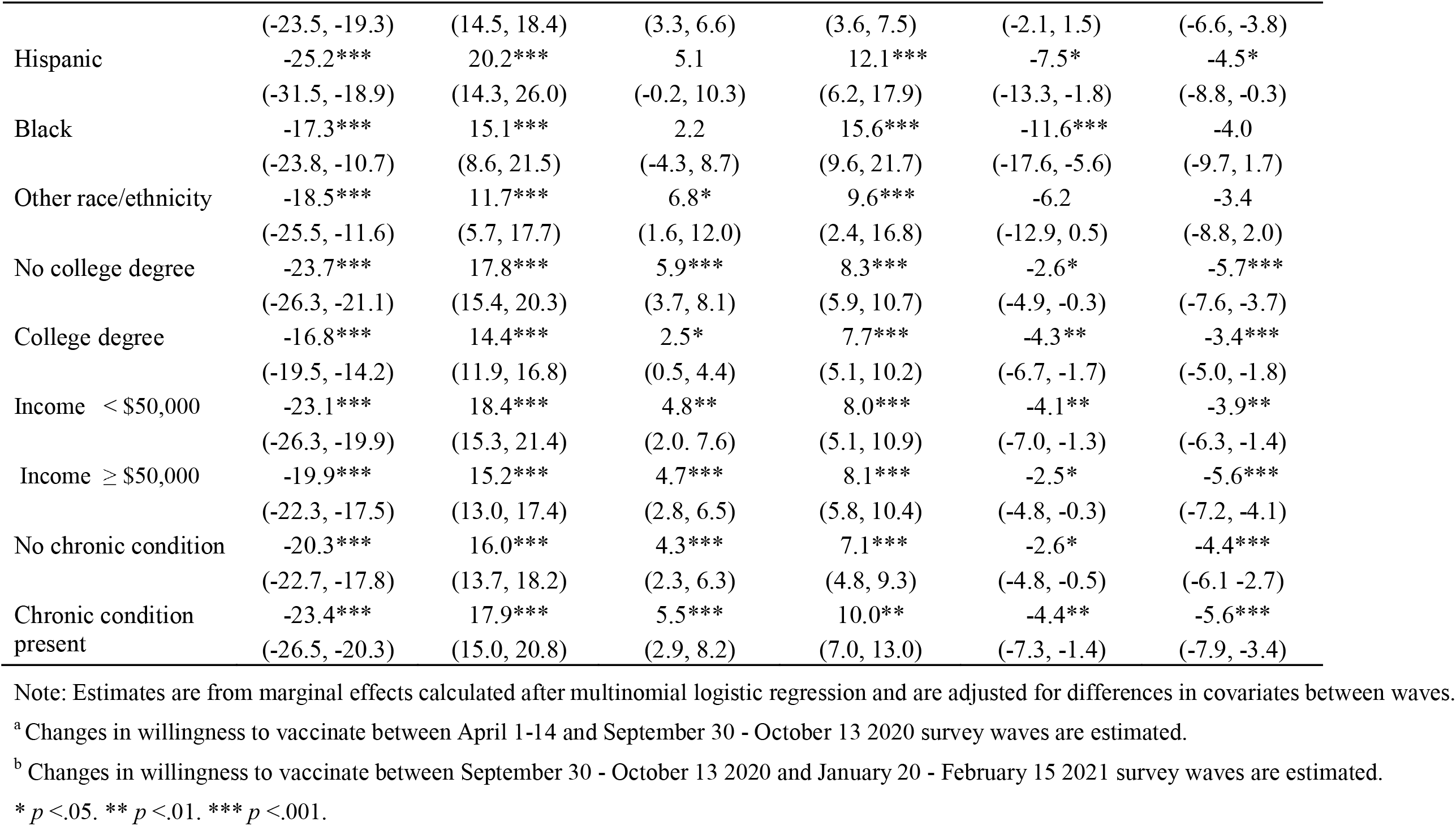
Regression estimates of percentage point changes in willingness to vaccinate between April and October 2020 and between October 2020 and February 2021 in the Understanding America Study (UAS).

|  | Changes in willingness to vaccinate |  |  | Changes in willingness to vaccinate |  |  |
| --- | --- | --- | --- | --- | --- | --- |
|  | between April and October 2020<sup>a</sup> |  |  | between October 2020 and February 2021<sup>b</sup> |  |  |
|  | Δ Willing to<br>accept vaccine | Δ Unwilling to<br>accept vaccine | Δ Undecided | Δ Willing to<br>accept vaccine | Δ Unwilling to<br>accept vaccine | Δ Undecided |
|  | %<br>(95% CI) | %<br>(95% CI) | %<br>(95% CI) | %<br>(95% CI) | %<br>(95% CI) | %<br>(95% CI) |
| Overall sample | -21.3***<br>(-23.3, -19.4) | 16.6***<br>(14.8, 18.4) | 4.7***<br>(3.1, 6.3) | 8.1***<br>(6.3, 9.9) | -3.2***<br>(-5.0, -1.5) | -4.9***<br>(-6.2, -3.5) |
| Age 18-39 | -22.0***<br>(-25.5, -18.4) | 17.4***<br>(14.1, 20.7) | 4.6**<br>(1.6, 7.5) | 5.2**<br>(1.8, 8.5) | -0.3<br>(-3.6, 2.9) | -4.8***<br>(-7.4, -2.2) |
| Age 40-59 | -23.9***<br>(-27.2, -20.5) | 18.5***<br>(15.4, 21.7) | 5.4***<br>(2.5, 8.2) | 10.5***<br>(7.5, 13.4) | -5.7***<br>(-8.5, -2.9) | -4.7***<br>(-7.1, -2.4) |
| Age 60+ | -18.1***<br>(-21.2, -15.1) | 13.8***<br>(11.0, 16.6) | 4.3***<br>(1.9, 6.7) | 9.2***<br>(6.3, 12.1) | -4.2**<br>(-7.0, -1.4) | -5.0***<br>(-7.1, -2.9) |
| Male | -20.3***<br>(-23.1, -17.6) | 15.1***<br>(12.5, 17.7) | 5.2***<br>(3.1, 7.4) | 6.5***<br>(3.8, 9.1) | -2.0<br>(-4.4, 0.5) | -4.5***<br>(-6.4, -2.6) |
| Female | -22.3***<br>(-25.0, -19.6) | 18.1***<br>(15.5, 20.6) | 4.2***<br>(1.8, 6.6) | 9.7***<br>(7.2, 12.1) | -4.4***<br>(-6.9, -2.0) | -5.2***<br>(-7.2, -3.2) |
| White | -21.4*** | 16.4*** | 4.9*** | 5.5*** | -0.3 | -5.2*** |
|  | (-23.5, -19.3) | (14.5, 18.4) | (3.3, 6.6) | (3.6, 7.5) | (-2.1, 1.5) | (-6.6, -3.8) |
| Hispanic | -25.2*** | 20.2*** | 5.1 | 12.1*** | -7.5* | -4.5* |
|  | (-31.5, -18.9) | (14.3, 26.0) | (-0.2, 10.3) | (6.2, 17.9) | (-13.3, -1.8) | (-8.8, -0.3) |
| Black | -17.3*** | 15.1*** | 2.2 | 15.6*** | -11.6*** | -4.0 |
|  | (-23.8, -10.7) | (8.6, 21.5) | (-4.3, 8.7) | (9.6, 21.7) | (-17.6, -5.6) | (-9.7, 1.7) |
| Other race/ethnicity | -18.5*** | 11.7*** | 6.8* | 9.6*** | -6.2 | -3.4 |
|  | (-25.5, -11.6) | (5.7, 17.7) | (1.6, 12.0) | (2.4, 16.8) | (-12.9, 0.5) | (-8.8, 2.0) |
| No college degree | -23.7*** | 17.8*** | 5.9*** | 8.3*** | -2.6* | -5.7*** |
|  | (-26.3, -21.1) | (15.4, 20.3) | (3.7, 8.1) | (5.9, 10.7) | (-4.9, -0.3) | (-7.6, -3.7) |
| College degree | -16.8*** | 14.4*** | 2.5* | 7.7*** | -4.3** | -3.4*** |
|  | (-19.5, -14.2) | (11.9, 16.8) | (0.5, 4.4) | (5.1, 10.2) | (-6.7, -1.7) | (-5.0, -1.8) |
| Income < \$50,000 | -23.1*** | 18.4*** | 4.8** | 8.0*** | -4.1** | -3.9** |
|  | (-26.3, -19.9) | (15.3, 21.4) | (2.0, 7.6) | (5.1, 10.9) | (-7.0, -1.3) | (-6.3, -1.4) |
| Income ≥ \$50,000 | -19.9*** | 15.2*** | 4.7*** | 8.1*** | -2.5* | -5.6*** |
|  | (-22.3, -17.5) | (13.0, 17.4) | (2.8, 6.5) | (5.8, 10.4) | (-4.8, -0.3) | (-7.2, -4.1) |
| No chronic condition | -20.3*** | 16.0*** | 4.3*** | 7.1*** | -2.6* | -4.4*** |
|  | (-22.7, -17.8) | (13.7, 18.2) | (2.3, 6.3) | (4.8, 9.3) | (-4.8, -0.5) | (-6.1, -2.7) |
| Chronic condition present | -23.4*** | 17.9*** | 5.5*** | 10.0** | -4.4** | -5.6*** |
|  | (-26.5, -20.3) | (15.0, 20.8) | (2.9, 8.2) | (7.0, 13.0) | (-7.3, -1.4) | (-7.9, -3.4) |

Note: Estimates are from marginal effects calculated after multinomial logistic regression and are adjusted for differences in covariates between waves.
<sup>a</sup> Changes in willingness to vaccinate between April 1-14 and September 30 - October 13 2020 survey waves are estimated.
<sup>b</sup> Changes in willingness to vaccinate between September 30 - October 13 2020 and January 20 - February 15 2021 survey waves are estimated.
\* $p < .05$ . \*\* $p < .01$ . \*\*\* $p < .001$ .

Intentions to vaccinate increased significantly by 8.1% (95% CI[6.3, 9.9], *p*<.001) from 52.7% to 60.8% between October 2020 and February 2021 (see Table 3). This was explained by a 4.9% (95% CI[3.5, 6.2], *p*<.001) decrease in the percentage of participants indicating they were undecided about accepting the COVID-19 vaccine and a 3.2% (95% CI[1.5, 5.0], *p*<.001) decline in the proportion of the population indicating they would refuse the vaccine from October 2020 to February 2021 (Table 3).

Our regression analyses showed statistically significant increases in intentions to vaccinate between October 2020 and February 2021 for all demographic groups examined (Table 3). Over this period the largest increases in willingness to vaccinate were found among Black (15.6% increase, 95% CI[9.6, 21.7], *p*<.001) and Hispanic participants (12.1% increase, 95% CI[6.2, 17.9], *p*<.001). Those aged 40-59 years (10.5% increase, 95% CI[7.5, 13.4], *p*<.001), those with chronic health conditions (10.0% increase, 95% CI[7.0, 13.0], *p*<.001), and females (9.7% increase, 95% CI[7.2, 12.1], *p*<.001) also showed relatively large increases in willingness to vaccinate, as shown in Table 3.

## 4. Discussion

This is the first nationally representative study to show a longitudinal increase in willingness to vaccinate since the emergence of evidence for the safety and efficacy of COVID-19 vaccines. After a rapid decline in intentions to vaccinate early in the pandemic,^5-7^ willingness to vaccinate increased by over eight percentage points from 52.7% to 60.8% among U.S. adults between October 2020 and February 2021. This was driven by a decline in the percentage of participants reporting being undecided about vaccination and a decrease in the percentage reporting they are unwilling to vaccinate. This trend is congruent with the findings of a recent U.S. study that used repeated cross-sectional surveys to show a decline in intentions to refuse the COVID-19 vaccine among priority groups between September and December 2020.^17^

A significant increase in willingness to vaccinate was observed across all demographic groups and was most pronounced among Black (15.6%) and Hispanic (12.1%) participants. This increase is important because COVID-19 vaccine acceptance has been particularly low among Black and Hispanic groups who are known to experience a disproportionate burden of severe illness and death due to COVID-19.^5,12,18^ As such, achieving a high levels of vaccine uptake among ethnic minority groups is crucial to reduce racial inequalities in disease outcomes due to COVID-19. However, despite the gains seen in recent months vaccine acceptance remained under 50% in Black participants in February 2021 and just above 60% in the overall sample. Furthermore, approximately one in three Black participants explicitly reported they will not be vaccinated (vaccine refusal) and this figure was similarly high among other population sub-groups (e.g. young adults, lower income and education groups). For this reason, public health messaging needs to continue to communicate the safety, efficacy and necessity of COVID-19 vaccines and to build trust in vaccines, particularly among Black populations.^12,19^

Understanding the drivers of changes in vaccination acceptance will now be critical in shedding light on beliefs and concerns that could be targeted to promote vaccination uptake and reduce loss of life due to the virus.^18^ Recent evidence suggests that there has been a decline in concern about the speed at which COVID-19 vaccines have been developed suggesting that the U.S. population has been at least partly assured that the science underpinning the vaccines and the vaccine approval process has not been rushed.^17,19^ However, public concerns about the safety and long-term health effects of the vaccine remain. For this reason, further effort needs to be invested in conveying accurate safety information through trusted sources (e.g. community groups, faith leaders, GPs) and tackling vaccine misinformation.^11,19-20^

The current study adds to the existing literature by examining national trends in willingness to vaccinate in a large-scale nationally representative sample of adults tracked continuously throughout the pandemic. However, the UAS is limited in that the survey is administered to community dwelling adults in English and Spanish only and is accessible only to those willing to engage with online surveys. This study is also limited in its reliance on measures of intentions to vaccinate and reported vaccination behavior.

In conclusion, this longitudinal nationally representative study found that willingness to vaccinate against COVID-19 has increased in the US from October 2020 to February 2021 and this increase has been largest in Black and Hispanic populations. However, almost 30% of the US population still intend not to be vaccinated and continued efforts to improve vaccine uptake are needed.

## Data Availability

NA

## Author Contributions

Dr Daly had full access to the study data and takes responsibility for the integrity of the data and accuracy of the data analysis.

### Concept and design

All authors.

### Acquisition, analysis, or interpretation of data

All authors.

### Drafting of the manuscript

All authors.

### Critical revision of the manuscript for important intellectual content

All authors.

### Statistical analysis

Daly.

## Acknowledgements

The project described in this paper relies on data from survey(s) administered by the Understanding America Study, which is maintained by the Center for Economic and Social Research (CESR) at the University of Southern California. The content of this paper is solely the responsibility of the authors and does not necessarily represent the official views of USC or UAS. The collection of the UAS COVID-19 tracking data is supported in part by the Bill & Melinda Gates Foundation and by grant U01AG054580 from the National Institute on Aging. However, these organizations bear no responsibility for the analysis or interpretation of the data. ER’s time was part-funded by the European Research Council and their support is gratefully acknowledged.

---

**Table S1.**
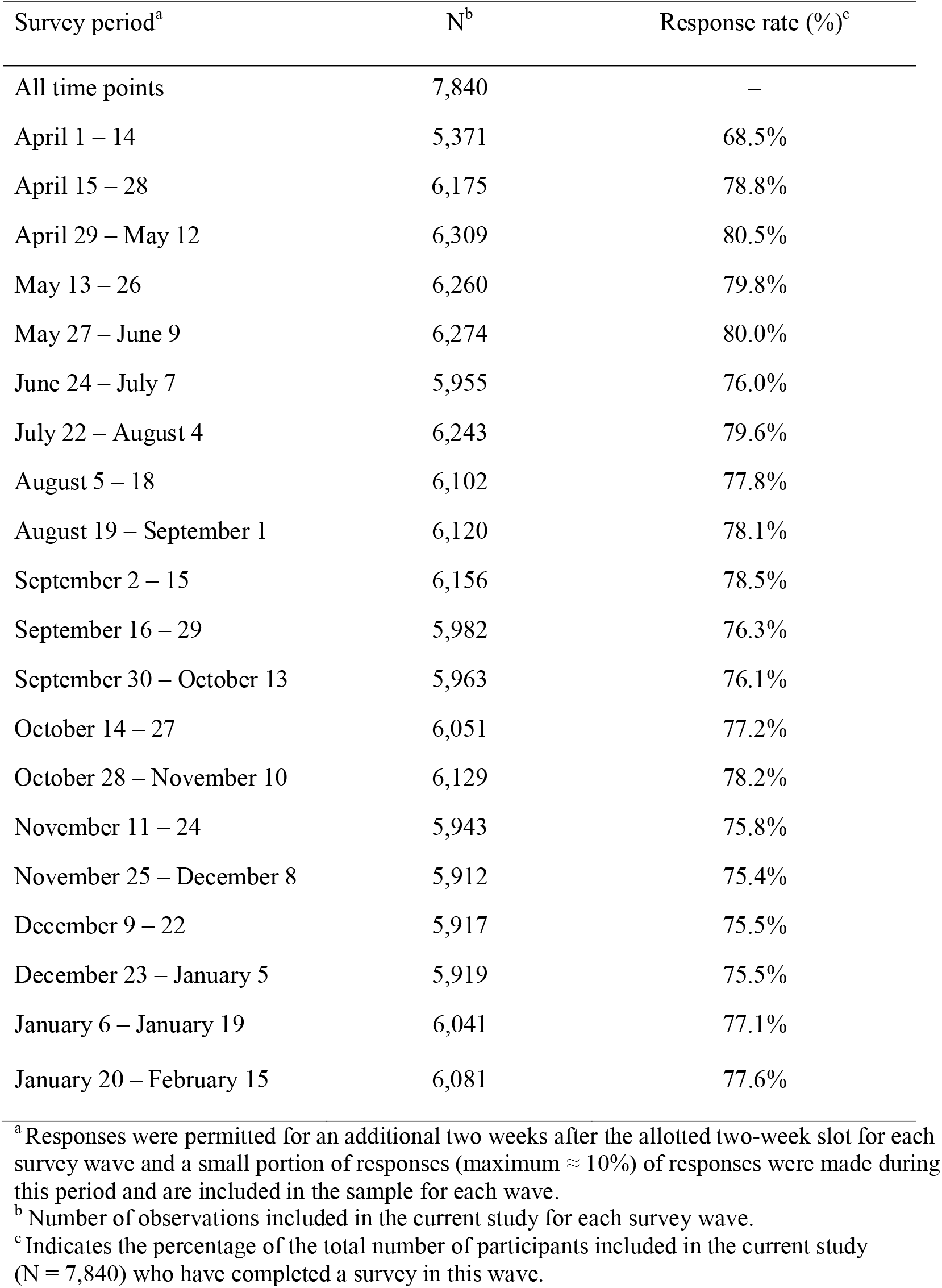
Number of participants and response rate among participants in the current study by survey wave from April 1 2020 to February 15 2021 (N = 7,840, Obs. = 120,903).

